# Comparing biomarkers for COVID-19 disease with commonly associated preexisting conditions and complications

**DOI:** 10.1101/2020.10.02.20205609

**Authors:** Jesse Huang

## Abstract

Severe coronavirus disease 2019 (COVID-19) has been associated with certain preexisting health conditions and can cause respiratory failure along with other multi-organ injuries. However, the mechanism of these relationships is unclear, and prognostic biomarkers for the disease and its systemic complications are lacking. This study aims to examine the plasma protein profile of COVID-19 patients and evaluate overlapping protein modules with biomarkers of common comorbidities.

Blood samples were collected from COVID-19 cases (n=307) and negative controls (n=78) among patients with acute respiratory distress. Proteins were measured by proximity extension assay utilizing next-generation sequencing technology. Its associations to COVID-19 disease characteristics were compared to that of preexisting conditions and established biomarkers for myocardial infarction (MI), stroke, hypertension, diabetes, smoking, and chronic kidney disease.

Several proteins were differentially expressed in COVID-19, including multiple pro-inflammatory cytokines such as IFN-γ, CXCL10, and CCL7/MCP-3. Elevated IL-6 was associated with increased severity, while baseline IL1RL1/ST2 levels were associated with a worse prognosis. Network analysis identified several protein modules associated with COVID-19 disease characteristics overlapping with processes of preexisting hypertension and impaired kidney function. BNP and NTpro-BNP, markers for MI and stroke, increased with disease progression and were positively associated with severity. MMP12 was similarly elevated and has been previously linked to smoking and inflammation in emphysema, along with increased cardiovascular disease risk.

In conclusion, this study provides an overview of the systemic effects of COVID-19 and candidate biomarkers for clinical assessment of disease progression and the risk of systemic complications.

## Introduction

The recent coronavirus (COVID-19) pandemic is caused by the severe acute respiratory syndrome coronavirus 2 (SARS-CoV2), and since its identification in late 2019, the virus has spread worldwide with over 30 million reported cases^1^. Clinical presentation can range from mild flu-like symptoms, including fever, cough, and shortness of breath to acute respiratory distress syndrome (ARDS) requiring mechanical ventilation^2-4^. Disease severity and mortality rate have been associated with certain preexisting conditions such as a history of cardiovascular disease, hypertension, diabetes, and obesity, which corresponds with worse prognosis^4-7^. These conditions often accompany older age and likely explain the higher mortality rate observed among the elderly population^4^. Recent genetic studies indicate the potential protective effect of specific blood antigens and possibly polymorphisms within the ACE2 gene^8,9^, the primary cell entry receptor for SARS-CoV2^10^.

Although clinical manifestations are mainly respiratory, early clinical reports and extrapolation from similar coronaviruses^11^ (e.g., SARS-CoV1 and MERS-CoV) have detailed the systemic effects of COVID-19, including acute cardiac injury, heart failure, arrhythmia, gastrointestinal distress, impaired liver function, and acute kidney injury^4,7,11-13^. Thromboembolic complications are common among patients with preexisting cardiac and cerebrovascular diseases, which is likely related to the systemic inflammation and pro-coagulatory conditions from COVID-19 infection^14,15^. Early surveillance studies have also reported neurological manifestations, including altered mental status and impaired consciousness along with fatigue, pain, and sensory disturbances (e.g., anosmia, dysgeusia) post-recovery^16,17^; however, the long-term complications of COVID-19 remain unclear.

While the clinical characteristics of COVID-19 are continually refined in real-time, more efficient tools, particularly prognostic markers, are needed to evaluate disease progression for targeted intervention strategies and better understand the overlapping systemic pathology between SARS-CoV2 infection and comorbidities. Using high-sensitivity proximity extension technology, this study examines the blood proteome of COVID-19 patients for protein markers associated with early infection and disease prognosis and compares with known biomarkers of common preexisting conditions and related complications.

## Methods

Adult patients (n=384) presenting with acute respiratory distress were investigated at the Massachusetts General Hospital (Boston, USA), of which 306 patients tested positive for COVID-19 while 78 remained as negative controls^18^. The descriptive statistics of the cohort are provided in **Table 1**. Longitudinal blood sampling was conducted for cases at 3, 7, and 28 days from baseline, if possible. Preexisting conditions including heart (e.g., coronary artery disease, congestive heart failure, valvular disease), lung (e.g., asthma, chronic obstructive pulmonary disease, regular O2 use), and kidney (e.g., chronic kidney disease, baseline creatinine >1.5) disease were recorded along with any history of diabetes, hypertension, and any immunocompromising conditions. Characterization of “obese” was defined as a body-mass index (BMI) of ≥30 kg/m^2^. The patient’s condition was assessed using a 6-point ordinal scale (1=death; 2=intubation and mechanical ventilation; 3= non-invasive ventilation or high-flow oxygen; 4=hospitalized with supplementary oxygen; 5=hospitalized without supplementary oxygen; 6=not hospitalized) based on World Health Organization guidelines^19^.

**Table 1.**
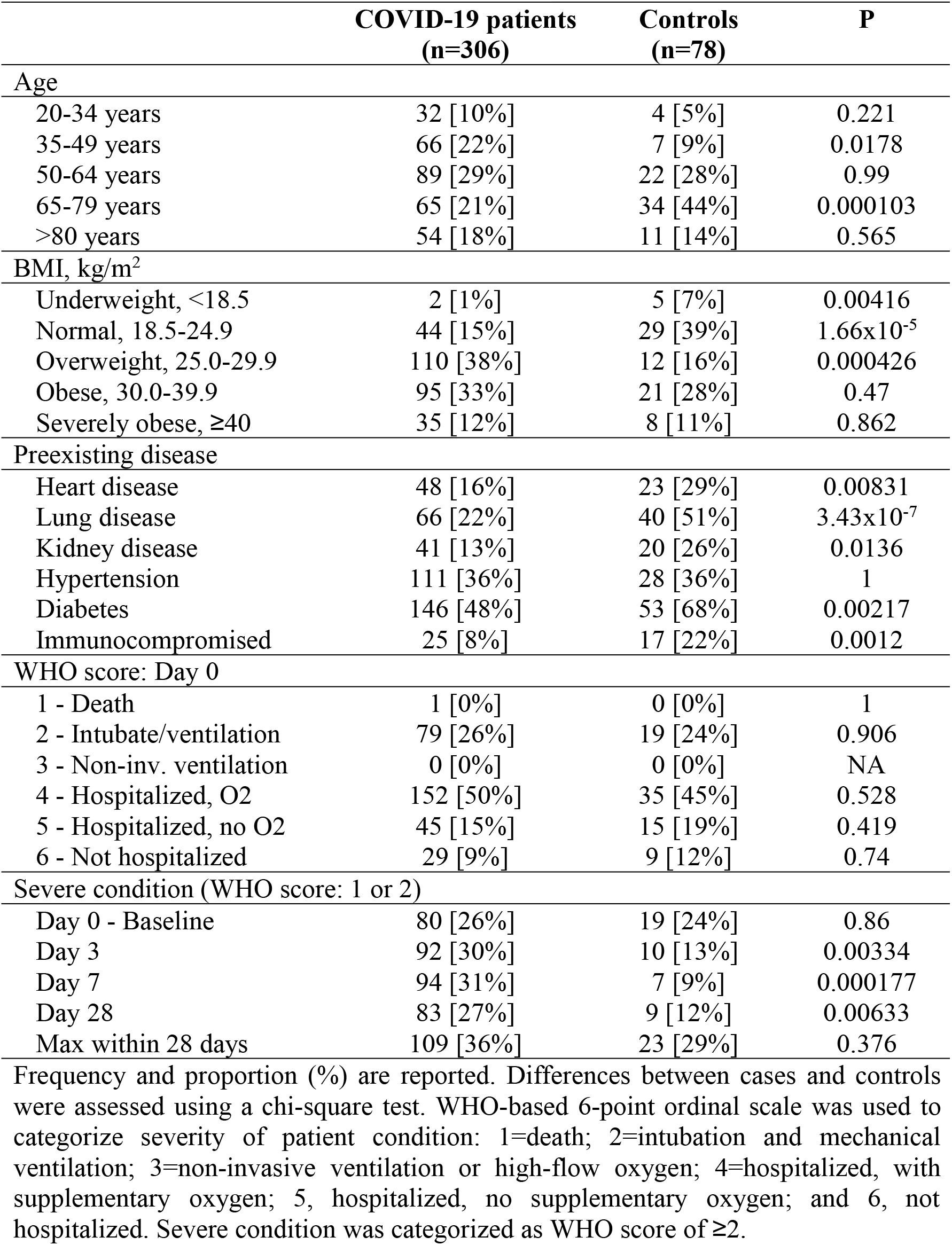
Descriptive statistics of the cohort.

Proteins were measured in plasma using a proximity extension assay (PEA), a high-sensitivity multiplex immunoassay that utilizes paired oligonucleotide antibody probes for protein identification followed by quantification using qPCR or next-generation sequencing (NGS) technology^20^. In this study, samples were analyzed with the NGS-based Olink Explore product consisting of four 384-plex panels of 1536 assay probes, including 48 controls and three inter-panel quality control markers (IL-6, IL-8, and TNF)^21^. The relative concentration for each protein was quantified as log base-two normalized protein expression (NPX) levels. Internal assay controls were used to quality check each step of the assay (i.e., incubation, extension, and amplification), and sample measures with high variability were excluded. Additional details regarding the method have been described elsewhere^20,21^.

In summary, 1420 unique proteins were analyzed with the majority having a call-rate (i.e., measurable levels above the limit of detection) above >80% (**Supplementary Table S1**). Measures below 25% (x=160) were excluded, while those between 25-80% (x=229) remained in the analysis but were interpreted with precaution. In total, 1260 proteins passed quality control for the analysis.

Differences in protein levels between COVID-19 positive patients and negative controls were analyzed using a multivariable linear regression model adjusting for age and preexisting conditions (Table 1). Longitudinal changes in protein concentrations were analyzed using a paired Student t-test. Preanalytical variation associated with sample quality was assessed and corrected using previously defined markers of sample handling (e.g., CD40L)^22^. Association with severity and prognosis was assessed using the baseline severity score or maximum-reached severity score within the 28 day period, respectively. Scores for severity and prognosis were dichotomized based on the usage of mechanical ventilation or death (severe: 1-2, non-severe, 3-6; WHO score). Significance after Bonferroni correction for multiple testing was set at P<10^−5^. All statistical analyses were conducted using R v.4.0.2 (Vienna, Austria).

Modules or clusters of proteins were identified by weighted correlation network analysis (WGCNA) using a Pearson-based weighted adjacency matrix (signed, β=14) and average linkage hierarchical clustering^23^. Modules were evaluated for enriched biological processes^24^ and used to compare study associations with the differential protein profiles of other diseases including myocardial infarction (MI)^25-27^, cardiovascular-related death (CVD) / heart failure (HF)^27^, stroke^27,28^, hypertension^29^, atherosclerosis^30^, diabetes^31^, smoking^32^, kidney function^33^, and chronic kidney disease (CKD)^33^.

## Results

Many proteins were differentially expressed among COVID-19 positive patients compared to negative controls, as shown in **Figure 1**. Inflammatory cytokines such as CXCL10, CXCL11, and IFN-γ increased four-fold at initial sampling but then decreased during the follow-up period. In contrast, lower levels were detected for CDON, ROR1, and BOC but likewise stabilized and regressed within the first week. However, several proteins, including ITGA11, continued to decrease over time. A delayed effect was observed with SDC1, PTN, and SFRP1, which were not associated initially but increased gradually as the disease progressed. Similarly, levels of ACE2 increased over two folds within the first week but was not elevated at baseline (β= −0.06, P= 0.65).

**Figure 1.**
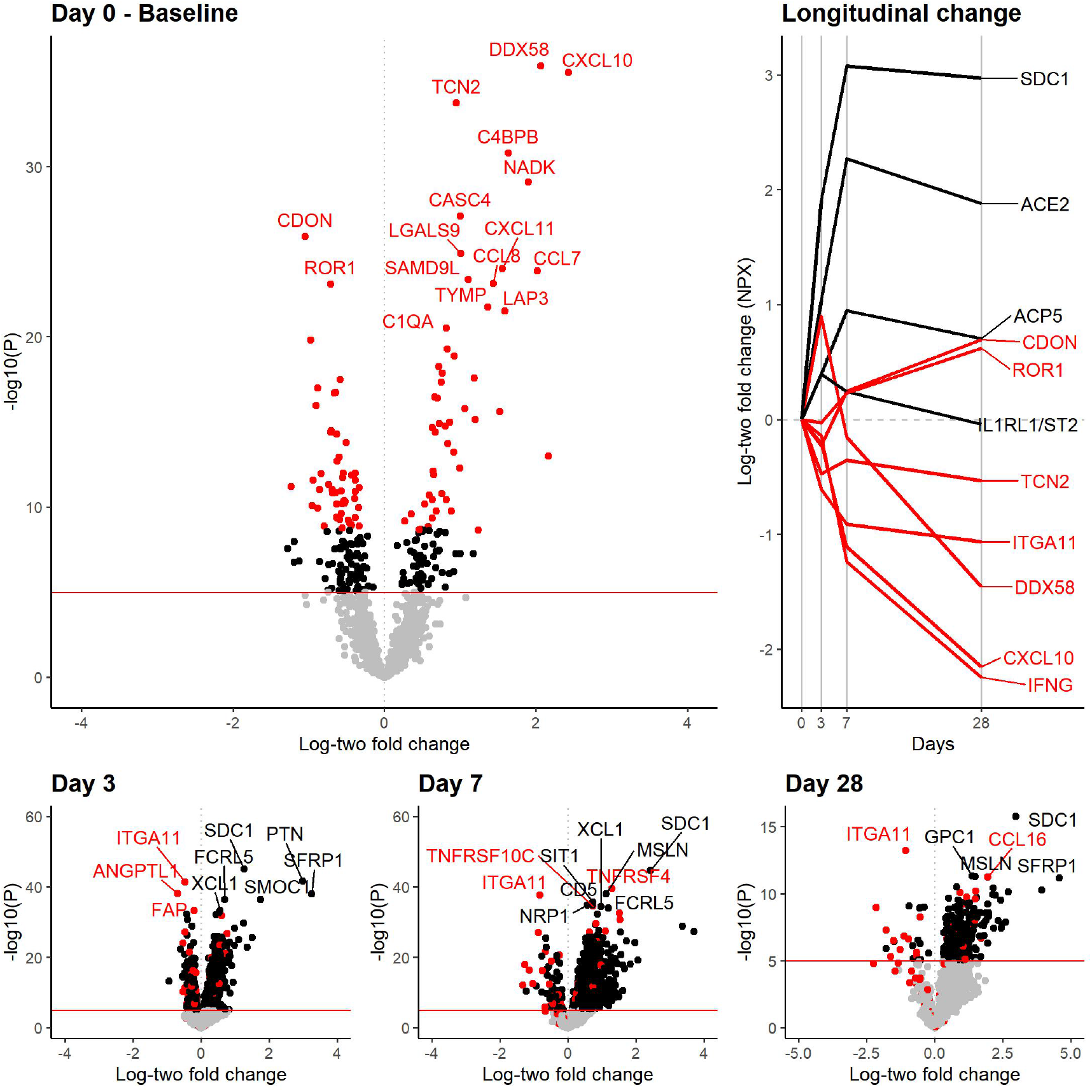
Proteins associated with COVID-19 and changes in their level during the disease course. Differences in log base-two protein levels (NPX) between COVID-19 positive cases (n=305) and negative controls (n=78) were analyzed using a multivariable linear regression model adjusting for age, preexisting conditions (see **Table 1**), and sample handling. The top one hundred COVID-19 associated markers are labeled in red for all plots. Changes in protein levels at 3, 7, and 28 days compared to baseline among cases (bottom panels) were analyzed using a paired Student t-test (n=211, 131, 40, respectively). Levels relative to baseline are also plotted longitudinally for those followed till day 28 (n=40) to limit potential survivorship bias.

Findings were not significantly affected after correcting for sample handling, although there was a minor improvement in the variability (**Supplementary Table S2, S3**). The majority remained significant after correcting for preexisting conditions and obesity. However, a few proteins, including IL1R2, a protein upregulated in the adipose tissue of obese patients^34^, were only associated after stratification by BMI (normal: β<0.001, P=0.99; obese β=-0.54, P=3.3×10^−7^; **Supplementary Table S4**).

Biomarkers associated with disease severity, as defined by mechanical ventilator use, are illustrated in **Figure 2** and compared to the disease-associated markers identified in **Figure 1**. Overlapping markers, including EZR, NADK, and KRT19, may be useful biomarkers for diagnosing infection and monitoring disease severity. IL-6, which was only slightly elevated among cases (β=0.60, P=0.02), was significantly correlated with severity. However, the majority of proteins associated with severity were different from those associated with the disease. Measures such as DDAH1 and NPM1 were also associated with severity among COVID-19 negative controls indicating a lack of specificity for specific proteins.

**Figure 2.**
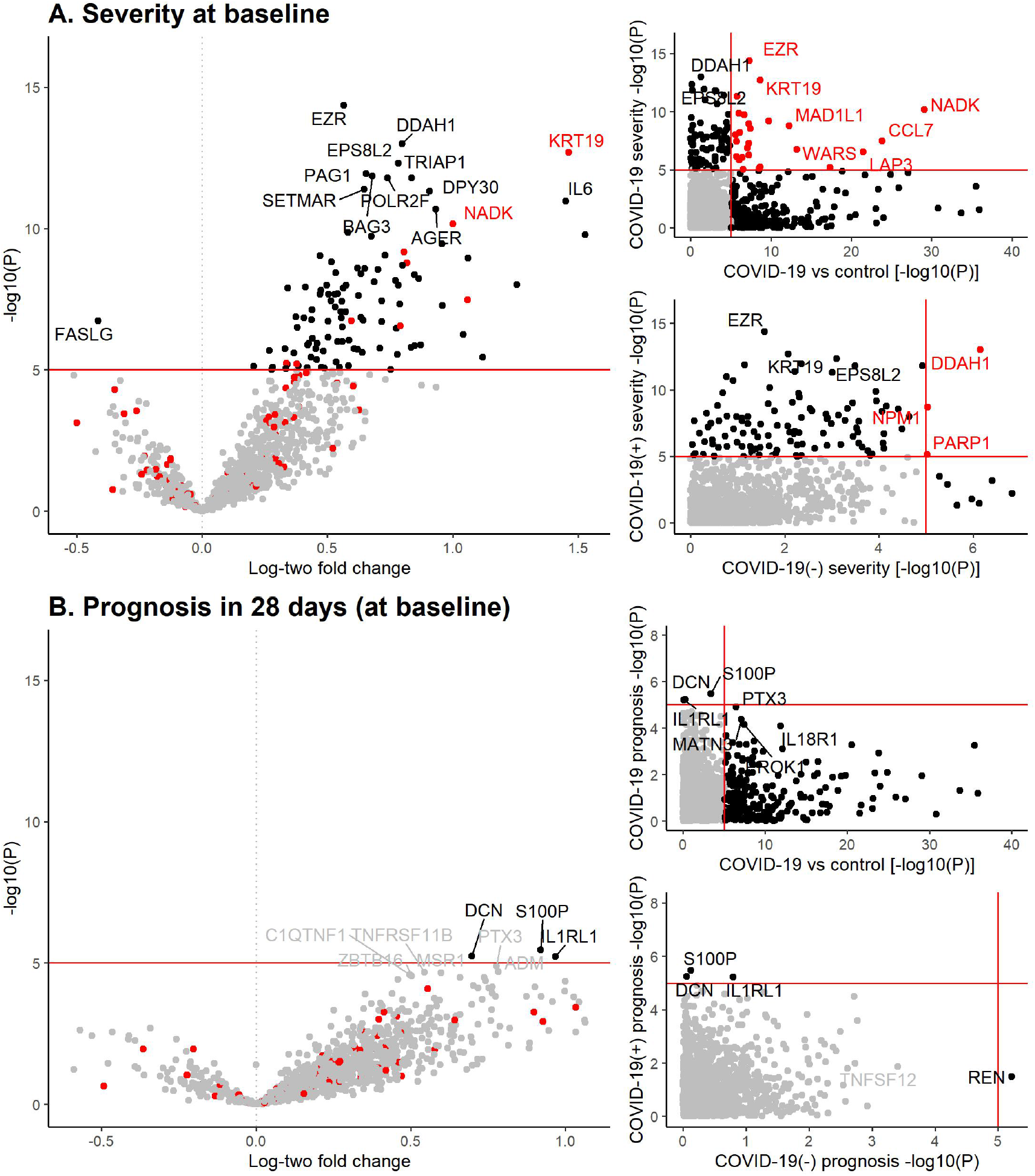
Proteins associated with COVID-19 severity and 28-day prognosis. Association between log base-two protein levels (NPX) measured at baseline and either [A] severity at baseline or [B] prognosis within 28 days was determined using a multivariable linear regression model adjusting for age, preexisiting conditions, sample handling, and baseline severity (for prognosis). Both severity and prognosis was defined as a WHO score of ≥2 (ie. Death or use of mechanical ventilation) at baseline or within 28 days, respectively.

Baseline levels of DCN, S100P, and the cardiac biomarker IL1RL1/ST2 were associated with maximum-observed severity (i.e., death or requiring ventilation) within the 28 days, even after correcting for baseline severity (**Supplementary Table S7**). Plasma levels of S100P were lower among cases compared to controls (β=-0.48, P=0.0004), but levels of DCN and IL1RL1 were not effected at baseline. However, IL1RL1 was associated with baseline severity (β=0.79, P=4.7×10^−8^). Chemokine CXCL10 was also correlated with baseline severity and prognosis (β=0.63, P=0.0003; β=0.90, P=0.0006).

As most cases were over 50 years old, many have preexisting conditions such as diabetes and hypertension (**Table 1**). COVID-19 positive patients were more likely to be obese but had unexpectedly lower rates of lung disease and diabetes, although this may be due to selection bias among controls. Association between baseline protein levels and preexisting conditions among cases was examined in **Figure 3**. Leptin (LEP), a metabolism-regulating hormone primarily secreted by adipose tissue^34^, was the primary protein associated with obesity but was only slightly increased among cases (β=0.681, P=0.0007). Cardiac biomarkers, NT-proBNP, and its active form NPPB/BNP, were negatively associated with obesity but positively associated with preexisting heart disease. Both were under-expressed in COVID-19 at baseline (β<-1.19, P<2×10^−7^) but increased significantly during follow up (β>1.1, P<3×10^−5^). BMI significantly modified baseline association for both cardiac biomarkers (normal: β>-0.71, P>0.20; obese: β<-1.82, P<2×10-8). FGF19 was elevated in patients with hypertension and possibly those with preexisting kidney disease (unadjusted: β=0.73, P=2×10^−5^; adjusted: β=0.77, P=0.08).

**Figure 3.**
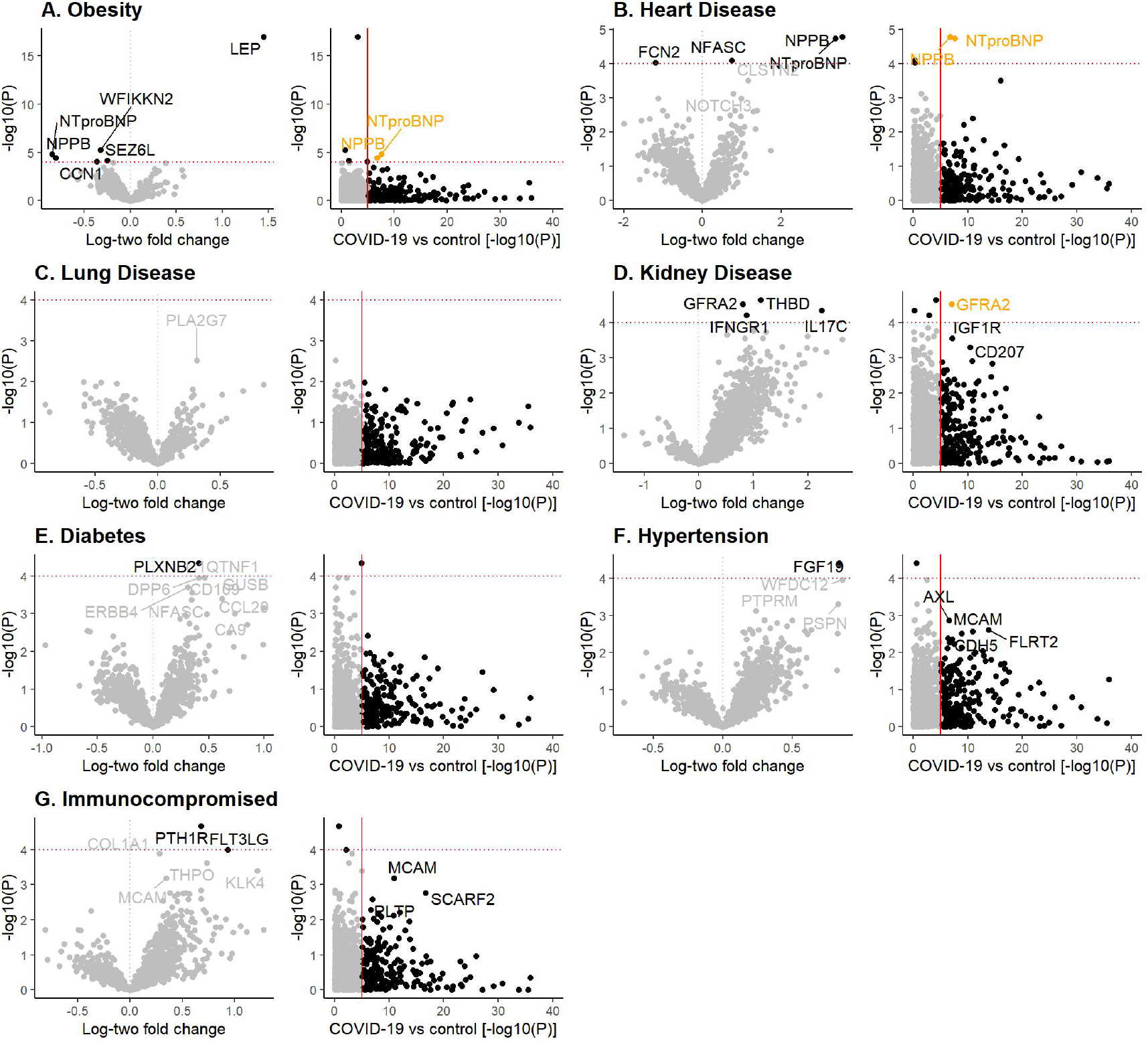
Proteins associated with preexisting conditions. Association between log base-two protein levels (NPX) and preexisting conditions including obesity (>30kg/m^2^), heart/lung/kidney disease, diabetes, hypertension, and any immunocompromising conditions are illustrated above (left) and in comparison to their association with COVID-19 (right; See **Figure 1**, baseline). Cases with preexisting conditions were compared to those with no preexisting condition (n=89) using a multivariable linear regression model adjusting for age and other preexisting conditions. Details of the analysis are available in **Supplementary Tables S8a-g**.

To determine if specific biological processes overlap between COVID-19 disease and related comorbidities, modules of intercorrelated proteins were identified and used to cross-examine associations between disease characteristics, preexisting conditions, and relevant biomarkers established in previous studies. As illustrated in **Figure 4**, several identified modules correspond to over- (green) and under- (turquoise/black) expressed proteins among COVID-19 patients. As previously illustrated, these tend to be different from proteins associated with disease progression (red/turquoise) and severity (yellow). There was a significant overlap in markers associated with preexisting kidney disease and hypertension (turquoise), which were responsive to COVID-19 infection and increased with disease progression. Many have been previously established as markers of reduced kidney function, based on estimated glomerular filtration rate (eGFR), and increased risk of developing chronic kidney disease.

**Figure 4.**
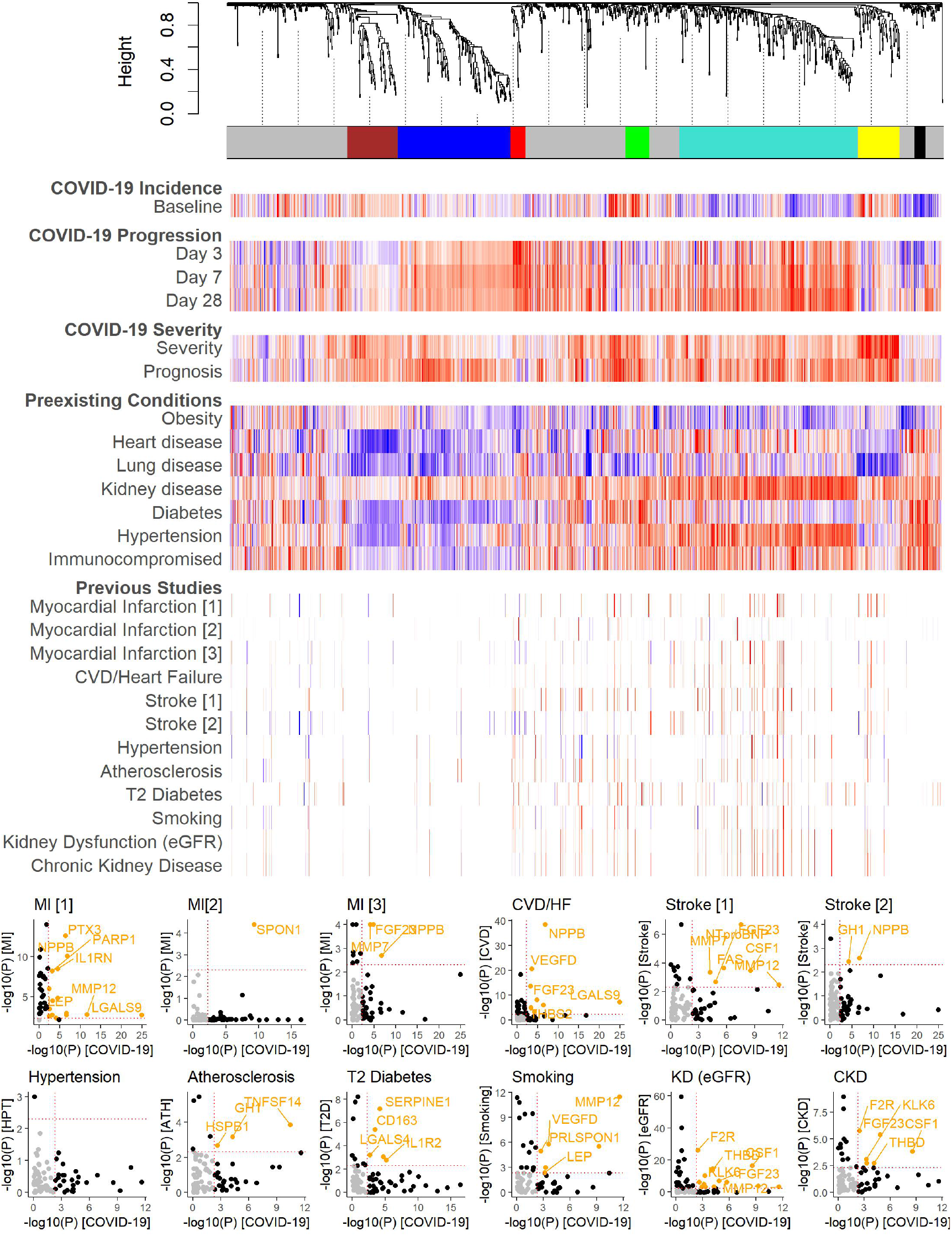
Comparison of protein modules associated with COVID-19 and disease-related preexisting conditions and complications. Dendrogram and colored bars illustrate intercorrelated protein modules among COVID-19 positive patients determined by weighted correlation network analysis (WGCNA). Colors indicate a normalized score (−1, blue; +1, red) of the effect estimate divided by standard error (SE) with color and intensity corresponding to the direction and strength of the association, respectively. Scores were normalized within each study by the 95th percentile of the absolute scores to allow inter-study comparability and minimize outlier effects. Studies are referenced in the methods by order. Additional details are provided in **Supplementary Table S9**.

NPPB and NT-proBNP (left-end turquoise) were associated with myocardial infarction, heart failure, and ischemic stroke. Its moderately correlated protein, MMP12, was similarly elevated in both myocardial infarction and stroke. MMP12 was also higher among smokers, being associated with inflammation in emphysema; therefore, it may be a shared proinflammatory mediator between COVID-19 and severity-related comorbidities. The protein module associated with disease severity (yellow) overlapped with biomarkers for increased myocardial infarction risk such as AGER, CTSL, PARP1, and SOD2. Many severity-related measures were also associated with preexisting lung disease, although in the opposite direction. Proteins of the last module (black), which includes ITGA11, continued to decrease throughout the 28 day observation period. Considering its moderate-overlap with obesity and diabetes, the module may be related to long-term metabolic dysfunctions.

## Discussion

This study provides a broad overview of the systemic effects of COVID-19 measured through blood, a natural sink for multiple organ systems and an easily accessible medium for clinical investigation. Findings indicate a significant disruption in the circulating proteome of infected patients, impacting multiple biological processes relating to pulmonary inflammation along with cardiac injury and renal dysfunction. These findings support the observation of multi-organ complications reported in previous clinical studies^4,7,36^.

As expected, many pro-inflammatory cytokines were elevated during the early stages of the disease and may influence overall severity, as shown in the previous studies^3,4^. The hyperactive immune response against SARS-CoV2 infection, often referred to as a cytokine storm, has been hypothesized to be a cause of disease mortality^37,38^. IFN-γ-induced chemoattractant CXCL10 was one of the primary cytokines elevated in cases and was a suggestive marker for disease severity. Previous studies have also shown increased levels of CXCL10 along with CCL7 (MCP-3) in COVID-19, and CXCL10 was a suggested biomarker for ARDS with protein DDX58^39,40^. As IL-6 blockade has been effective for managing cytokine release syndrome, IL-6 has also been investigated as a therapeutic target for COVID-19^37^. Although not notably elevated among cases in this study, higher levels of IL6 were associated with increased severity. Findings further support the potential benefits of such treatment. As IL-6 is also a marker with myocardial infarction and smoking, treatment efficacy may be modulated by active cigarette smoking or preexisting cardiovascular disease targeting related disease complications^25,27^.

Several cardiac biomarkers were influenced by COVID-19, including the hormone brain natriuretic peptide (BNP/NPPB) and its inactivated-form, N-terminal pro–BNP (NT-proBNP). Both are known predictors of acute cardiac injury and heart failure^27,41^ and have been proposed as measures for increased mortality among COVID-19 patients^42^. A retrospective examination of deceased COVID-19 patients has also shown elevated levels of circulating NT-proBNP during hospitalization^4^. Therefore, BNP-related measures may be useful for monitoring cardiac stress and the risk of thromboembolic complications, particularly among those with obese or preexisting heart conditions. Another biomarker, IL1RL1, also known as ST2, is associated with cardiac remodeling, and soluble ST2 has been a marker of acute myocardial infarction^43-45^. Elevated levels of ST2 was associated with increased mortality rate and may be an additional complementing marker of cardiac complications for COVID-19^46^.

Other proteins may also indicate cardiac-related injury, including CDON, which along with its coexpressed partner BOC, were lower among cases. CDON deficiency in mice has been associated with cardiac remodeling and fibrosis through hyperactivation of the Wnt/β-catenin pathway and may indicate an increased risk of cardiac injury and heart failure in patients^47^. CDON levels were also negatively correlated with severity and, to a lesser extent, disease prognosis. On the other hand, MMP12, an increased measure associated of cigarette smoking, maybe a mediator of pulmonary inflammation^32,48^, in COVID-19 and preexisting lung conditions, and cardiovascular-related comorbidities.

As the primary entry receptor for SARS-CoV2, angiotensin-converting enzyme 2 (ACE2), an essential moderator of blood pressure, has often been examined in the relationship between COVID-19 and cardiovascular complications. Studies have suggested that coronavirus infection can affect the expression of ACE2 pathways in the heart and increase cardiac complication risk associated with localized inflammation^49^. ACE inhibitors frequently used to manage hypertension have been associated with increased risk of kidney injury among COVID-19 patients^50^. Furthermore, increased levels of soluble ACE2, as seen among cases in this study, may offer some protection against SARS-CoV2 infection by inhibiting receptor-binding activity, although this requires further clinical investigation^51^.

Studies have also shown ACE2 expression in kidneys, which may indicate a direct relationship between SARS-CoV2 infection and renal complications^52^. Systematic release of pro-inflammatory cytokines in ARDS may increase the risk of acute kidney injury, while its resulting accumulation from reduced renal function may, in turn, exacerbate ARDS^53^. In this study, several proteins were associated with both COVID-19 and kidney dysfunction, and maybe possible biomarkers for monitoring acute injury, particularly among those with hypertension.

The long-term complication of COVID-19 could not be directly assessed in this study due to the limited follow-up time. However, previous studies have reported lasting changes in metabolic and sensory functions, particularly among severe cases^17,35^. Although levels of proteins like ILTGA11 seemed to show long-term disruptions from disease, proper longitudinal studies will be required to investigate its trajectory. Although we focused on circulating proteins in this study, other media such as sputum may provide more localized measures for COVID-19^48^. Similarly, urine and cerebrospinal fluid may be better for assessing renal and neurological complications. However, blood remains the single most comprehensive source for assessing systemic effects, and the sensitivity of proximity extension technology allows the detection of trace proteins from multiple organ systems. Unfortunately, previous studies have been limited to targeting specific aspects of the blood proteome, which significantly limits the resolution for comparing overlapping biological processes. However, the recent incorporation of NGS readout in PEA will likely provide a more comprehensive disease proteomic profile in future studies.

In summary, this study provides an initial assessment of the overlapping biological processes associated with COVID-19 and related comorbidities. Cardiac biomarkers NTpro-BNP, BNP, and ST2, may be useful for monitoring and assessing the risk of cardiac and cerebrovascular complications while specific measures of inflammation such as CXCL10, IL6, and MMP12 may be useful for identifying patient groups responsive to immunosuppressive treatments. However, further investigations are required to validate the efficacy of these potential biomarkers in clinical settings.

## Data Availability

Study data is available for request as of October 2, 2020.

https://info.olink.com/mgh-covid-study-overview-page

## Abbreviations

ACE2: Angiotensin-converting enzyme 2
ARDS: Acute respiratory distress syndrome
BMI: Body mass index
CVD: Cardiovascular-related death
CKD: Chronic kidney disease
COVID-19: Coronavirus disease 2019
eGFR: Estimated glomerular filtration rate
HF: Heart failure
IL-6: Interleukin-6
IL1RL1/ST2: Interleukin 1 receptor-like 1
LEP: Leptin
MERS: Middle Eastern respiratory syndrome
MI: Myocardial infarction
NGS: Next-generation sequencing
NPPB/BNP: B-type natriuretic peptide
NPX: Normalized protein expression
NT-proBNP: N-terminal pro–B-type natriuretic peptide
PEA: Proximity extension assay
SARS: Severe acute respiratory syndrome
SARS-CoV2: Severe acute respiratory syndrome coronavirus 2
WGCNA: Weighted gene correlation network analysis

## Acknowledgments

The author would like to thank the principal investigators of the study (Michael R. Filbin, Alexandra-Chloe Villani, Nir Hacohen, Marcia Goldberg) for providing the accessible clinical and proteomic data which was processed in collaboration with Olink. Additional thanks to the collection team (Kyle Kays, Kendall Lavin-Parsons, lair Parry, Brendan Lilley, Carl Lodenstein, Brenna McKaig, Nicole Charland, Hargun Khanna, Justin Margolin); processing team (Moshe Sade-Feldman, Anna Gonye, Irena Gushterova, Tom Lasalle, Nihaarika Sharma, Brian C. Russo, Maricarmen Rojas-Lopez, Kasidet Manakongtreecheep, Jessica Tantivit, Molly Fisher Thomas); and those involved with data preprocessing (Arnav Mehta, Alexis Schneider)^18^.

## References

1. Dong E, Du H, Gardner L. An interactive web-based dashboard to track COVID-19 in real time. Lancet Infect Dis. 2020;20(5):533–4.

2. Wang D, Hu B, Hu C, et al. Clinical Characteristics of 138 Hospitalized Patients With 2019 Novel Coronavirus-Infected Pneumonia in Wuhan, China. JAMA. 2020;323(11):1061–9.

3. Huang C, Wang Y, Li X, et al. Clinical features of patients infected with 2019 novel coronavirus in Wuhan, China. Lancet. 2020;395(10223):497–506.

4. Chen T, Wu D, Chen H, et al. Clinical characteristics of 113 deceased patients with coronavirus disease 2019: retrospective study. BMJ. 2020;368:m1091.

5. Ko JY, Danielson ML, Town M, et al. Risk Factors for COVID-19-associated hospitalization: COVID-19-Associated Hospitalization Surveillance Network and Behavioral Risk Factor Surveillance System. Clin Infect Dis. 10.1093/cid/ciaa1419

6. Yang J, Zheng Y, Gou X, et al. Prevalence of comorbidities and its effects in patients infected with SARS-CoV-2: a systematic review and meta-analysis. Int J Infect Dis. 2020;94:91–5.

7. Kochi AN, Tagliari AP, Forleo GB, Fassini GM, Tondo C. Cardiac and arrhythmic complications in patients with COVID-19. J Cardiovasc Electrophysiol. 2020;31(5):1003–8.

8. Ellinghaus D, Degenhardt F, Bujanda L, et al. Genomewide Association Study of Severe Covid-19 with Respiratory Failure. N Engl J Med. 10.1056/NEJMoa2020283

9. Hou Y, Zhao J, Martin W, et al. New insights into genetic susceptibility of COVID-19: an ACE2 and TMPRSS2 polymorphism analysis. BMC Med. 2020;18(1):216.

10. Hoffmann M, Kleine-Weber H, Schroeder S, et al. SARS-CoV-2 Cell Entry Depends on ACE2 and TMPRSS2 and Is Blocked by a Clinically Proven Protease Inhibitor. Cell. 2020;181(2):271-280.e8.

11. de Wit E, van Doremalen N, Falzarano D, Munster VJ. SARS and MERS: recent insights into emerging coronaviruses. Nat Rev Microbiol. 2016;14(8):523–34.

12. Gupta A, Madhavan MV, Sehgal K, et al. Extrapulmonary manifestations of COVID-19. Nat Med. 2020;26(7):1017–32.

13. Docherty A, Harrison E, Green C, et al. Features of 16,749 hospitalised UK patients with COVID-19 using the ISARIC WHO Clinical Characterisation Protocol. medRxiv 2020.04.23.20076042

14. Lodigiani C, Iapichino G, Carenzo L, et al. Venous and arterial thromboembolic complications in COVID-19 patients admitted to an academic hospital in Milan, Italy. Thromb Res. 2020;191:9–14.

15. Klok FA, Kruip MJHA, van der Meer NJM, et al. Incidence of thrombotic complications in critically ill ICU patients with COVID-19. Thromb Res. 2020;191:145–7.

16. Varatharaj A, Thomas N, Ellul MA, et al. Neurological and neuropsychiatric complications of COVID-19 in 153 patients: a UK-wide surveillance study. Lancet Psychiatry. 10.1016/S2215-0366(20)30287-X

17. Halpin SJ, McIvor C, Whyatt G, et al. Postdischarge symptoms and rehabilitation needs in survivors of COVID-19 infection: A cross-sectional evaluation. J Med Virol. 10.1002/jmv.26368

18. Filbin MR, Villani AC, Hacohen N, and Goldberg M. MGH COVID-19 study:Collaborative effort to investigate the plasma proteomic signatures of COVID-19 positive patients. Olink, Sep. 11 2020 (accessed Sep. 11, 2020)

19. WHO. COVID-19 therapeutic trial synopsis. Geneva: World Health Organization, Feb 18, 2020. https://www.who.int/docs/default-source/blue-print/covid-19-therapeutic-trial-synopsis.pdf (accessed Sep. 15, 2020).

20. Assarsson E, Lundberg M, Holmquist G, et al. Homogenous 96-plex PEA immunoassay exhibiting high sensitivity, specificity, and excellent scalability. PLoS ONE. 2014;9(4):e95192.

21. Olink. PEA – a high-multiplex immunoassay technology with qPCR or NGS readout. v1.0, May 26, 2020. https://www.olinkexplore.com/content/uploads/2020/09/olink-white-paper-pea-on-ngs-qpcr-v1.0.pdf (accessed Sep. 16, 2020).

22. Shen Q, Björkesten J, Galli J, et al. Strong impact on plasma protein profiles by precentrifugation delay but not by repeated freeze-thaw cycles, as analyzed using multiplex proximity extension assays. Clin Chem Lab Med. 2018;56(4):582–594.

23. Langfelder P, Horvath S. WGCNA: an R package for weighted correlation network analysis. BMC Bioinformatics. 2008;9:559.

24. Subramanian A, Tamayo P, Mootha VK, et al. Gene set enrichment analysis: a knowledge-based approach for interpreting genome-wide expression profiles. Proc Natl Acad Sci USA. 2005;102(43):15545–50.

25. Gidlöf O, Evander M, Rezeli M, Marko-Varga G, Laurell T, Erlinge D. Proteomic profiling of extracellular vesicles reveals additional diagnostic biomarkers for myocardial infarction compared to plasma alone. Sci Rep. 2019;9(1):8991.

26. Kulasingam A, Hvas AM, Grove EL, Funck KL, Kristensen SD. Detection of biomarkers using a novel proximity extension assay in patients with ST-elevation myocardial infarction. Thromb Res. 2018;172:21–8.

27. Ferreira JP, Sharma A, Mehta C, et al. Multi-proteomic approach to predict specific cardiovascular events in patients with diabetes and myocardial infarction: findings from the EXAMINE trial. Clin Res Cardiol. 10.1007/s00392-020-01729-3

28. Lind L, Siegbahn A, Lindahl B, Stenemo M, Sundström J, Ärnlöv J. Discovery of New Risk Markers for Ischemic Stroke Using a Novel Targeted Proteomics Chip. Stroke. 2015;46(12):3340–7.

29. Lin YT, Fall T, Hammar U, et al. Proteomic Analysis of Longitudinal Changes in Blood Pressure. J Clin Med. 2019;8(10):E1585.

30. Lind L, Ärnlöv J, Lindahl B, Siegbahn A, Sundström J, Ingelsson E. Use of a proximity extension assay proteomics chip to discover new biomarkers for human atherosclerosis. Atherosclerosis. 2015;242(1):205–10.

31. Molvin J, Pareek M, Jujic A, et al. Using a Targeted Proteomics Chip to Explore Pathophysiological Pathways for Incident Diabetes-The Malmö Preventive Project. Sci Rep. 2019;9(1):272.

32. Huang B, Svensson P, Ärnlöv J, Sundström J, Lind L, Ingelsson E. Effects of cigarette smoking on cardiovascular-related protein profiles in two community-based cohort studies. Atherosclerosis. 2016;254:52–8.

33. Carlsson AC, Ingelsson E, Sundström J, et al. Use of Proteomics To Investigate Kidney Function Decline over 5 Years. Clin J Am Soc Nephrol. 2017;12(8):1226–35.

34. Meier CA, Bobbioni E, Gabay C, Assimacopoulos-Jeannet F, Golay A, Dayer JM. IL-1 receptor antagonist serum levels are increased in human obesity: a possible link to the resistance to leptin. J Clin Endocrinol Metab. 2002;87(3):1184–8.

35. Wu Q, Zhou L, Sun X, Yan Z, Hu C, Wu J, Xu L, Li X, Liu H, Yin P, Li K, Zhao J, Li Y, Wang X, Li Y, Zhang Q, Xu G, Chen H. Altered lipid metabolism in recovered SARS patients twelve years after infection. Sci Rep 2017;7:9110.

36. Gavriatopoulou M, Korompoki E, Fotiou D, et al. Organ-specific manifestations of COVID-19 infection. Clin Exp Med. 10.1007/s10238-020-00648-x

37. Henderson LA, Canna SW, Schulert GS, et al. On the Alert for Cytokine Storm: Immunopathology in COVID-19. Arthritis Rheumatol. 2020;72(7):1059–63.

38. Cao X. COVID-19: immunopathology and its implications for therapy. Nat Rev Immunol. 2020;20(5):269–70.

39. Yang Y, Shen C, Li J, et al. Plasma IP-10 and MCP-3 levels are highly associated with disease severity and predict the progression of COVID-19. J Allergy Clin Immunol. 2020;146(1):119-127.e4.

40. Xie Y, Liu K, Luo J, et al. Identification of DDX58 and CXCL10 as Potential Biomarkers in Acute Respiratory Distress Syndrome. DNA Cell Biol. 2019;38(12):1444–51.

41. Weber M, Hamm C. Role of B-type natriuretic peptide (BNP) and NT-proBNP in clinical routine. Heart. 2006;92(6):843–9.

42. Qin JJ, Cheng X, Zhou F, et al. Redefining Cardiac Biomarkers in Predicting Mortality of Inpatients With COVID-19. Hypertension. 2020;76(4):1104–12.

43. Kakkar R, Lee RT. The IL-33/ST2 pathway: therapeutic target and novel biomarker. Nat Rev Drug Discov. 2008;7(10):827–40.

44. Shimpo M, Morrow DA, Weinberg EO, et al. Serum levels of the interleukin-1 receptor family member ST2 predict mortality and clinical outcome in acute myocardial infarction. Circulation. 2004;109(18):2186–90.

45. Jónsdóttir B, Ziebell Severinsen M, von Wowern F, San Miguel C, Goetze JP, Melander O. ST2 Predicts Mortality In Patients With Acute Hypercapnic Respiratory Failure Treated With Noninvasive Positive Pressure Ventilation. Int J Chron Obstruct Pulmon Dis. 2019;14:2385–93.

46. Pascual-Figal DA, Manzano-Fernández S, Boronat M, et al. Soluble ST2, high-sensitivity troponin T- and N-terminal pro-B-type natriuretic peptide: complementary role for risk stratification in acutely decompensated heart failure. Eur J Heart Fail. 2011;13(7):718–25.

47. Jeong MH, Kim HJ, Pyun JH, et al. Cdon deficiency causes cardiac remodeling through hyperactivation of WNT/β-catenin signaling. Proc Natl Acad Sci USA. 2017;114(8):E1345–E1354.

48. Babusyte A, Stravinskaite K, Jeroch J, Lötvall J, Sakalauskas R, Sitkauskiene B. Patterns of airway inflammation and MMP-12 expression in smokers and ex-smokers with COPD. Respir Res. 2007;8:81.

49. Oudit GY, Kassiri Z, Jiang C, Liu PP, Poutanen SM, Penninger JM, Butany J. SARS-coronavirus modulation of myocardial ACE2 expression and inflammation in patients with SARS. Eur J Clin Invest 2009;39:618–625.

50. Oussalah A, Gleye S, Clerc Urmes I, et al. Long-Term ACE Inhibitor/ARB Use Is Associated with Severe Renal Dysfunction and Acute Kidney Injury in Patients with severe COVID-19: Results from a Referral Center Cohort in the North East of France. Clin Infect Dis. 10.1093/cid/ciaa677

51. Hofmann H, Geier M, Marzi A, et al. Susceptibility to SARS coronavirus S protein-driven infection correlates with expression of angiotensin converting enzyme 2 and infection can be blocked by soluble receptor. Biochem Biophys Res Commun. 2004;319(4):1216–21.

52. Hamming I, Timens W, Bulthuis ML, Lely AT, Navis G, van Goor H. Tissue distribution of ACE2 protein, the functional receptor for SARS coronavirus. A first step in understanding SARS pathogenesis. J Pathol. 2004;203(2):631–7.

53. Joannidis M, Forni LG, Klein SJ, et al. Lung-kidney interactions in critically ill patients: consensus report of the Acute Disease Quality Initiative (ADQI) 21 Workgroup. Intensive Care Med. 2020;46(4):654–72.

